# Agreement Between Self- and Caregiver-Report of Thought Disturbances in Adults with Williams Syndrome and Intellectual Disability

**DOI:** 10.64898/2025.12.15.25341842

**Authors:** Sarah G Vassall, Andrew R Kittleson, Annalise S Halverson, Gabriella V Schock, Essence Leslie, Elisabeth Dykens, Elizabeth Roof, Julia M Sheffield, Kimberly S Bress

**Author notes:** Essence Leslie now affiliated with Department of Psychiatry, University of Pittsburgh Medical Center, 230 Mckee Place, Suite 138, Pittsburgh, PA, 15213, USA. **Corresponding Author:** Kimberly S Bress, Department of Psychiatry and Behavioral Sciences, Vanderbilt University Medical Center, Vanderbilt Psychiatric Hospital, 1601 23rd Avenue South, Suite 3050, Nashville, Tennessee, 37212, USA.

## Abstract

Williams Syndrome (WS) is a rare neurodevelopmental disorder associated with intellectual disability and increased vulnerability to traits such as anxiety, perseveration, and belief inflexibility. In the general population, these traits are linked to self-reported thought disturbances such as paranoia and delusions. However, little is known about how such disturbances present in WS, largely due to concerns regarding the validity of self-report in this population. To address this gap, we collected self- and caregiver-reported measures characterizing thought disturbances and related cognitive traits in adults with WS, assessing inter-rater reliability and correlations among measures. Total scores were similar across reporters, except for delusional ideation, which participants endorsed more strongly than caregivers. Several participants also reported clinically significant levels of paranoia, delusions, and worry that were not captured by caregiver report. These findings suggest that self-report is a valid method for accurately characterizing the severity and nature of thought disturbances in WS.

## 1 Background

Williams Syndrome (WS) is a neurodevelopmental disorder caused by deletion of genes on chromosome 7 and is associated with multisystem effects, including intellectual disability (Korenberg et al., 2000; Morris, 1993; Pober, 2010) ranging from profound to minimal (Fisher et al., 2016; Miezah et al., 2020). Deficits are most pronounced in complex cognitive domains such as working memory, attention, and executive functioning (Miezah et al., 2020). Beyond general cognitive differences, many individuals with WS also exhibit a characteristic social-affective profile marked by atypical social insight, elevated anxiety, perseveration or rumination, and belief inflexibility (Järvinen et al., 2013; Morel et al., 2018).

In the general population, these specific traits—differences in cognition (Freeman, 2016; Freeman et al., 2008), atypical social experiences (Carvalho et al., 2017; Misiak, 2025), worry (Freeman et al., 2020, 2012; Freeman and Garety, 2014), anxiety, (Sheffield et al., 2021; Therman et al., 2014; Thewissen et al., 2011) and belief inflexibility (Bronstein et al., 2019; Linney et al., 1998)—are linked to elevated risk for disturbances of thought, including paranoia and delusions (Garety et al., 2005; Freeman et al., 2002; Freeman, 2006; Bell and O’Driscoll, 2018). Such disturbances occur on a continuum, presenting with varying degrees of subclinical severity in an estimated 10-15% of individuals (see Freeman, 2007 for a review). Because the WS phenotype encompasses many of the risk factors for paranoia and delusions, examining the incidence of thought disturbances in this population is of particular interest. Such disturbances may exacerbate existing cognitive and processing difficulties, increase vulnerability to additional psychiatric diagnoses, and hinder the effectiveness of therapeutic or occupational interventions. However, very little is known about the extent to which individuals with WS experience paranoia or delusions; to our knowledge, there are no studies to date that have explicitly and systematically measured the incidence and severity of thought disturbances in WS.

This knowledge gap is perpetuated in large part by lack of consensus regarding how best to characterize paranoia, delusions, and other thought disturbances in individuals with intellectual disabilities (Finlay and Lyons, 2001). Self-report is a common, if not cornerstone, methodology for evaluating and monitoring outcomes across clinical and research settings in psychiatry, particularly for disturbances of thought, perception, and mood. However, the administration and interpretation of self-report in populations with intellectual disabilities can be complicated by impairments in language, attention, working memory, executive function, and other cognitive domains, raising concerns regarding accuracy and reliability, even in adulthood (Balboni et al., 2013). These concerns are compounded by response biases to which individuals with intellectual disabilities are particularly vulnerable, including acquiescence, naysaying, latency, and recency biases (Emerson et al., 2013). Furthermore, many self-report measures of psychiatric symptomatology are not specifically validated for use in individuals with intellectual disabilities. As a result, caregiver or informant reports are often used as a proxy for self-reports for these individuals.

Studies of self-report reliability in individuals with intellectual disabilities are limited and often concern quality of life (QoL) reporting. The results of these studies are mixed, with some reporting high agreement between self- and informant-reports (Balboni et al., 2013; Berástegui et al., 2021; Santoro et al., 2022; Schmidt et al., 2010) and others reporting poor agreement (Janssen et al., 2005; Koch et al., 2015; Zimmermann and Endermann, 2008). Importantly, some QoL studies indicate that there is a higher degree of concordance for measures of physical quality of life compared to emotional or psychological measures (Berástegui et al., 2021; Santoro et al., 2022; Schmidt et al., 2010). Thus, agreement between self and caregiver-report is likely to vary greatly based on the nature of the report, as externalized behaviors (e.g., aggression, conduct disturbance, hyperactivity) may be more easily and accurately perceived by a caregiver compared to disturbances of thought or mood (e.g., paranoia, delusions, worry). Self-caregiver agreement is also improved when the individual with intellectual disability is older (Finlay and Lyons, 2001; Sheldrick et al., 2012), has a higher intelligence quotient (IQ), and demonstrates stronger and social communication abilities (Kaat and Lecavalier, 2015).

Characterizing the incidence and severity of thought disturbances in WS is necessary to fully understand the cognitive and psychological profile of this population. This is particularly important given the unique risk factors for thought disturbances which are common to the WS phenotype. Given the internal (i.e., unobservable) nature of thought disturbances, self-report is generally preferable to proxy-report when measuring these experiences. However, as there are well-documented challenges and limitations to collecting self-report in individuals with intellectual disabilities, the most appropriate method to measure thought disturbances in WS is unclear. To date, we are unaware of any work investigating the reliability and validity of self-reported thought disturbances in this population. Thus, the goals of this study are to characterize thought disturbances and related cognitive traits in adults with WS, and to investigate the degree to which self-report of thought disturbances can be reliably and validly collected in adults with WS. This validation is a necessary precedent to future research seeking to characterize how thought disturbances may manifest in WS, and interact with cognitive, social, and functional outcomes.

Using a battery of self- and caregiver-reported measures of thought disturbances (e.g., paranoia, delusions) and associated cognitive-affective features (e.g., anxiety, perseveration, cognitive insight), we provide a preliminary profile of these psychopathologies in adults with WS and investigate the degree of alignment between self- and caregiver-reports. Additionally, we elucidate how age, cognitive ability, and adaptive functioning influence the agreement between self- and caregiver-report. Ultimately, this work provides guidance for future clinical and research activities characterizing psychiatric symptomatology in individuals with intellectual disabilities broadly, a population which is at high risk for psychiatric and behavioral disturbances while being systematically underserved.

## 2 Methods

### 2.1 Participant Recruitment & Data Acquisition

We recruited a sample of 20 adults with a confirmed diagnosis of Williams Syndrome (WS) and their caregivers to complete a battery of self-report measures and cognitive assessments (Section 2.3, **Supplementary Table 1**). All procedures were approved by the Institutional Review Board for human subjects and conducted in accordance with the guidelines and regulations on ethical human research set forth in the Declaration of Helsinki; all participants and caregivers provided informed assent and consent, respectively, prior to participation in any study activities.

Participants and their caregivers completed one virtual visit via secure video call, and one in-person data collection session. During the virtual visit, each participant met with a member of the study team (KSB, EL) without their caregiver to complete the battery of self-report measures. To mitigate individual differences in literacy and comprehension, each question and all answer choices were read aloud for every measure, with the administrator recording the answer choice selected by the participant. Participants who could not successfully complete the example questions were excluded from further participation *(N=*1). In line with guidance concerning interviewing individuals with intellectual disabilities (McDonald et al., 2022), the administrator, when necessary, tailored self-reports to use plain language (i.e., no jargon), provided clarification of complex vocabulary, or rephrased questions with complex structures (example accommodations provided in **Supplementary Table 2**).

Caregivers asynchronously completed the same battery of self-report measures via REDCap (https://projectredcap.org), a secure virtual survey platform, blind to the responses provided by the participant. In addition, they completed a brief demographic and psychiatric history and the Vineland Adaptive Behavior Scale, 3^rd^ Edition (VABS-3; Sparrow et al., 2016) to evaluate the participant’s adaptive functioning.

During the in-person data collection session, held 2-4 weeks after the virtual visit, participants completed the Kaufman Brief Intelligence Test, 2^nd^ Edition (KBIT-2; Kaufman & Kaufman, 2013), evaluating nonverbal (NVIQ) and verbal (VIQ) intelligence. Additionally, if a self-report measure could not be completed during the virtual visit due to pacing or fatigue, that measure was administered during the in-person visit using the guided approach described above.

### 2.2 Participant Sample

Of the 20 individuals with Williams Syndrome recruited, 19 (*N* male=11; ages 18-56 years, *x̄*=32.42±9.11) demonstrated comprehension of the self-report instructions and successfully completed all measures (**Table 1**). Of the 19 associated caregivers, 18 were parents (17 mothers, 1 father) and one was an adult sibling (brother). All participants and their caregivers were white, with non-Hispanic ethnicity. Per caregiver report, none of the participants had ever been diagnosed with a schizophrenia-spectrum disorder (e.g., schizophrenia, schizotypal personality disorder, schizophreniform disorder, schizoaffective disorder, delusional disorder, brief psychotic disorder) or had ever been hospitalized for a psychiatric indication. However, two participants had been diagnosed with psychiatric conditions without psychotic features (panic disorder, n=1; anxiety and depression, n=1).

**Table 1.** Participant Sample Characterization. The demographic and cognitive characteristics of the participant sample are shown, including results of the Kauffman Brief Intelligence Test – Second Edition (KBIT-2), administered to the participant, and the Vineland Adaptive Behavior Scales – Third Edition (VABS-3), completed by the caregiver. We also report whether the participant was living with or separate from the caregiver, and in what setting. *Prior reports of rate of independent or low-support living in adults with WS ranges from 0% to about 13%% (Davies et al., 1998; Elison et al., 2010; Fisher et al., 2016).

| **Participants with Wiliams Syndrome (N=19, 8 females)** |  |  |
| --- | --- | --- |
| *Demographics* |  |  |
|  | *Mean ± SD* | *Range* |
| Age (Years) | 32.42±9.11 | 18-56 |
| *Cognitive Scores* |  |  |
|  | *Mean ± SD* | *Range* |
| KBIT-2 Composite | 66.79 ± 16.02 | 42-94 |
| KBIT-2 Nonverbal | 69.74 ± 17.75 | 40-98 |
| KBIT-2 Verbal | 71.16 ± 14.19 | 40-104 |
| VABS-3 Composite | 57 ± 10.56 | 37-77 |
| VABS-3 Communication | 49.06 ± 22.8 | 21-96 |
| VABS-3 Daily Living | 59.59 ± 9.98 | 40-71 |
| VABS-3 Socialization | 65.56 ± 8.83 | 45-80 |
| *Living Arrangement\** |  |  |
|  | *N* | *Percent* |
| With parent caregiver | 15 | 78% |
| With sibling caregiver | 1 | 6% |
| Group home | 1 | 6% |
| Independent with support | 2 | 10% |

As noted in **Table 1**, participants differed in their living arrangements (living with caregiver: 15 (parent), 1 (adult sibling); living independently: 3; **Figure 1L**) as well as their occupational status (full-time student: 2; part-time employment: 3; full-time employment: 3; unemployed nonstudent: 10; **Figure 1L**). Of those not employed, the primary daily activity reported by the caregiver included structure day programs (4), volunteering or community service (2), or activities at home (4).

**Figure 1.**
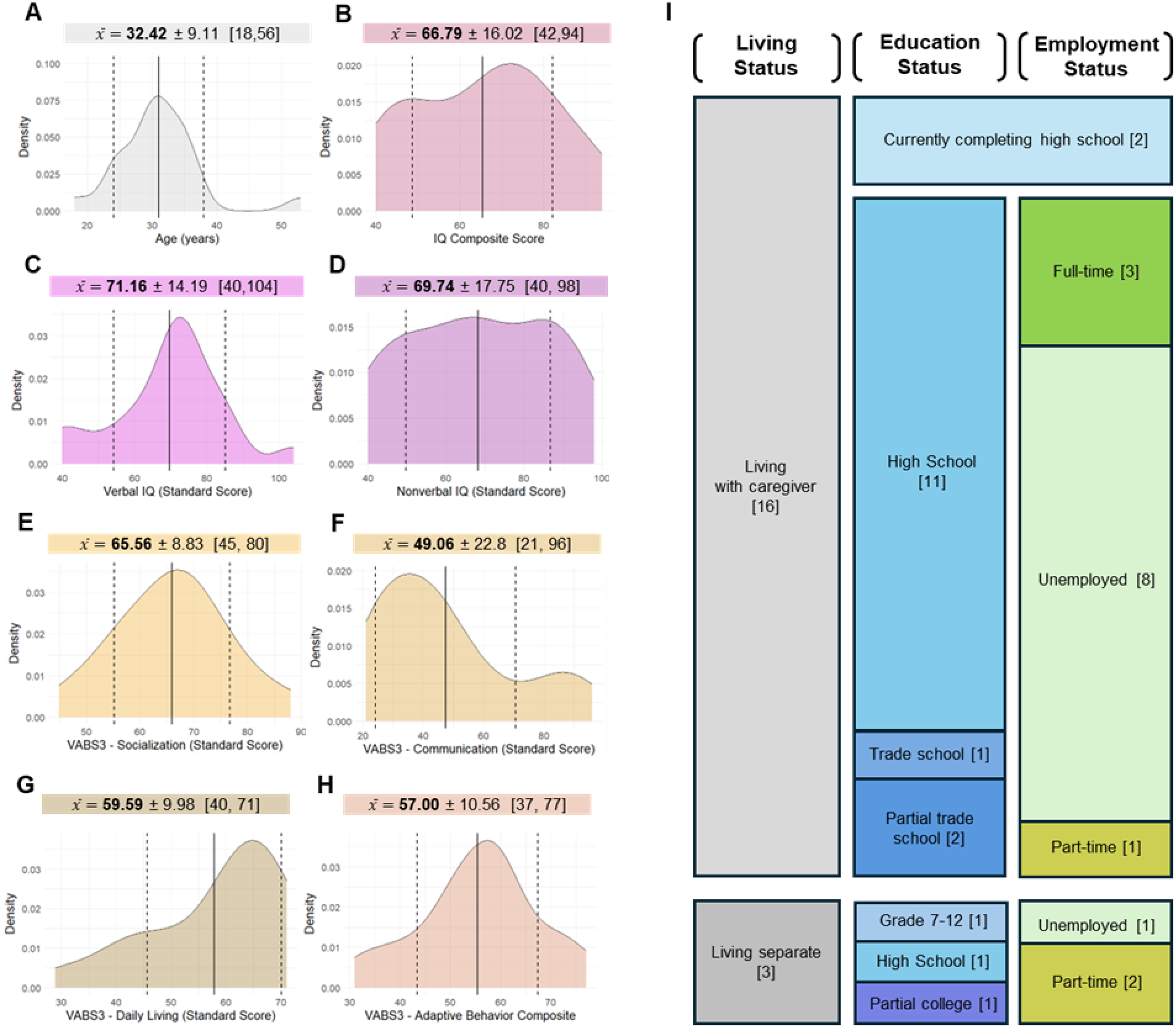
Demographic and Clinical Variable Distributions. **A.** Age distribution of the participant sample. **B-D.** Distributions of IQ composite (**B**), verbal IQ (**C**), and nonverbal IQ (**D**) standard scores. **E-H.** Distributions of VABS3 social domain (**E**), communication domain (**F**), daily living domain (**G**), and the adaptive behavior composite (**H**) standard scores. **I.** Proportions of participants living with or separate from caregivers (‘Living arrangement’), proportions of participants by highest level of education completed (‘Education status’), and proportion of employment status (‘Employment status’). The number of participants is shown in brackets within each box.

As measured by the KBIT-2, the mean composite IQ was 66.79±16.02 (**Figure 1B**), with a mean VIQ of 71.16±14.19 (**Figure 1C**) and mean NVIQ of 69.74±17.75 (**Figure 1D**). Adaptive functioning in the domains of socialization (65.56±8.83; **Figure 1E**), communication (49.06±22.8; **Figure 1I),** and daily living (59.59±9.98; **Figure 1J),** as well as overall adaptive functioning (57±10.56; **Figure 1K**) were measured using the VABS-3.

### 2.3 Self- and Caregiver-Reported Measures

Participants and their caregivers completed the following battery of report measures. In the current study, all measures have been normed on healthy individuals without a history of psychiatric illness. Except for the Difficulties in Emotion Regulation Scale (DERS) and Lubben Social Network Scale (LSNS), all measures were also normed on a clinical sample. All measures show strong internal consistency (**Supplementary Table 3**). No sex differences have been reported for these scales. It should be noted that none of these scales have been normed in populations with intellectual disabilities. These measures were specifically chosen to characterize either explicit thought disturbances (e.g., paranoia) or known associated risk factors (e.g., cognitive insight). The purpose and structure of each measure is described in detail below:

- **Revised-Green Paranoid Thoughts Scale** (r-GPTS; Freeman et al., 2021) assesses paranoid thinking. It consists of two 8-item subscales: GPTSa assesses ideas of reference (e.g., “people have been dropping hints for me”) and GPTSb assesses persecutory ideation (e.g., “I was convinced there was a conspiracy against me”). Items are scored on a 0-4 Likert scale, with higher scores indicating elevated paranoia endorsement (Min Total: 0; Max Total: 72). 16 items.
- **The Peters Delusion Inventory 21** (PDI-21; Peters et al., 2004) is designed to evaluate the presence and nature of unusual beliefs, as well as the distress, preoccupation, and conviction associated with these beliefs. Respondents state whether they endorse an unusual belief with a yes or no response (e.g., “Do you ever think people can communicate telepathically”) (Min Total: 0; Max Total: 21). 21 items.
- **The Dunn Worry Questionnaire** (DWQ; Freeman et al., 2020) consists of 2 subscales that assess general worry and paranoid worry. The 10-item general worry scale reflects the impact of overall worry (e.g., “worry has stopped me focusing on important things in my day”) while the 5-item paranoid worry scale reflects specific worries about harm inflicted by others (e.g., “anything and everything has set my mind thinking about people trying to upset me”). Items are scored on a 0-4 Likert scale, with higher scores indicating more severe impact of worry (Min Total: 0, Max Total: 60). 15 items.
- **The Liebowitz Social Anxiety Scale** (LSAS; Liebowitz, 1987). The LSAS assesses the degree of anxiety individuals experience in social situations. The LSAS consists of two 24-item subscales assessing: 1) the fear or anxiety induced by a certain situation (e.g., making eye contact, giving a presentation), and 2) how often the person avoids that situation. Items are scored on a 0-3 Likert scale, with higher scores indicating more severe fear and more frequent avoidance, respectively (Min Total (subscale): 0; Max Total (subscale): 72). 48 items.
- **The Perseverative Thinking Questionnaire** (PTQ; Ehring et al., 2011) assesses repetitive negative thinking, particularly worry and rumination (e.g., “I get stuck on certain issues and can’t move on”). Items are scored on a 0-4 Likert scale, with higher scores indicating more severe perseverative thinking or rumination (Min Total: 0; Max Total: 60). 15 items.
- **The Difficulties with Emotion Regulation Scale-16** (DERS-16; Gratz & Roemer, 2004) captures consequences of feeling upset and difficulties managing emotions (e.g., “when I am upset, I feel out of control”). Items are scored on a 1-5 Likert scale, with higher scores indicating more frequent incidence of emotional dysregulation and related externalizing behaviors (Min total: 16; Max total: 80). 16 items.
- **The Beck Cognitive Insight Scale** (BCIS; Beck et al., 2004) was developed specifically for clinical populations to measure not only patients’ conviction of their anomalous cognitive experiences (self-certainty), but also their ability to recognize “specific misinterpretations” (i.e., recognize that the cognitive experiences are anomalous; self-reflectiveness). The BCIS thus consists of two subscales: a 9-item scale of self-reflectiveness (SR) and a 6-item scale of self-certainty (SC). Items are scored on a 0-3 Likert scale, with greater difference between the subscale scores (SR-SC) reflecting greater cognitive insight (Min Total: 0 [SR], 0 [SC]; Max Total: 27 [SR], 18 [SC]). 15 items.
- **The Lubben Social Network Scale Abbreviated** (LSNS-6; Lubben et al., 2006) was designed to evaluate an individual’s perceived degree of social support, and was normed on 7,432 elderly adults. The 6-item measure is scored on a 0-5 Likert scale, with a higher score indicating a larger quantity and quality of individuals in one’s social support network (Min Total: 0, Max Total: 30). 6 items.
- **The Davos Assessment of Cognitive Biases Scale** (DACOBS; van der Gaag et al., 2013) was designed to measure cognitive biases, cognitive limitations, and safety behaviors by asking individuals about their thought patterns and beliefs across different bias categories. We collected three DACOBS subscales, assessing the biases of jumping to conclusions (JTC), belief inflexibility (BIF), and social cognition problems (SOC). Each 6-item subscale is scored on a 1-7 Likert scale, with a higher score indicating greater endorsement of the bias (Min Total (subscale): 6; Max Total (subscale): 42). 16 items.

### 2.4 Hypotheses and Statistical Approach

The statistical approach for each specific hypothesis is described in detail below. All statistical analyses were conducted in R (R Core Team, 2017). Linear regregssion and mixed effects models used in these analyses were fitted using the *lme4* package (Bates et al., 2015), with restricted maximum likelihood (REML; Corbeil & Searle, 1976). Significance testing for fixed effects was conducted using t-tests with Satterthwaite’s approximation for degrees of freedom, and estimated marginal means (EMMs) were computed using the *emmeans* package (Lenth, 2017).

#### 2.4.1 Between-Measure Correlations

To validate that the total scores were moderately (R=0.4-0.7) or strongly (R=0.7-1) intercorrelated as expected from general population patterns (e.g., associations between worry, perseveration, and paranoia; (Freeman et al., 2012)), we computed Pearson’s correlation coefficients (R) between all measures within each group using the *rcorr* function from the *Hmisc* package.

#### 2.4.2 Participant-Caregiver Total Score Agreement

We next sought to understand how well participants and their caregivers aligned on total scores for each measure. We hypothesized that there would be more participant-caregiver agreement for measures of observable behaviors (e.g., emotion regulation, socialization, etc.) and less agreement for measures of internal processes (e.g., paranoia, worry). In line with prior findings, we also hypothesized that older individuals and individuals with higher IQ scores would exhibit greater total score agreement with their caregiver.

For each measure, we used a paired Wilcoxon sign rank test to evaluate systematic differences between participant and caregiver scores, correcting for multiple comparisons using FDR. We then calculated the Pearson’s correlation coefficient (R) between participant-caregiver total scores for each measure. This approach assesses how closely participant and caregiver total scores co-vary across individuals, capturing consistency in scores rather than the mean differences or discrepancies.

Next, because the measures differ in their scoring ranges and maximum possible differences, we standardized participant-caregiver differences across measures by computing the percent of maximum possible difference (POMPD). Specifically, POMPD was defined as the absolute difference between the participant and caregiver total score divided by the maximum possible difference (maximum possible score minus minimum possible score). We then fit a linear mixed effects model on POMPD, including measure, composite IQ, and age as fixed effects and participant-caregiver pair as a random effect (i.e., random slope; **Model 1**):

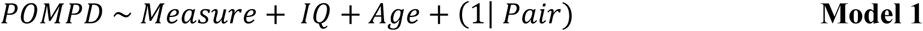

#### 2.4.3 Participant-Caregiver Item-Level Agreement

In addition to agreement in total scores, we also assessed participant-caregiver agreement at the level of individual measure items. For each measure, we estimated intraclass correlations (ICCs) across all items between each participant–caregiver pair using the *irr* package (Shrout & Fleiss, 1979) (two–way random effects, single measurement, absolute agreement; ICC[2,1]). We hypothesized that ICCs would be higher for measures with more items capturing externalizing behaviors compared to measures more concerned with internal (i.e., emotional or psychological) experiences.

To test whether agreement differed across measures, we additionally computed normalized difference in item-level responses (NDIR) as the absolute difference between the WS item response and caregiver item response, divided by the maximum possible difference for that item. As ICC is skewed by the size of the Likert scale, this approach normalizes the item-level differences across measures to allow for comparison. We then fit linear mixed–effects models with NDIR as the outcome, measure, IQ and age as fixed effects, and participant–caregiver pair as a random effect (**Model 2**). Pairwise comparisons between measures were then performed using estimated marginal means (*emmeans*) with FDR correction.

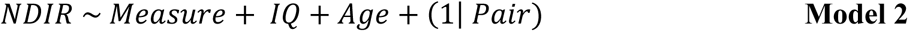

#### 2.4.4 Clinical Thresholds

Finally, given that many self-report measures are used clinically as well as in research settings, we conducted an exploratory analysis to understand whether the outcome of meeting or exceeding clinical thresholds differed across participants and their caregivers. This was conducted for measures with a validated clinical threshold, including the r-GPTS (18; (Freeman et al., 2021), PDI-21 (8; Peters et al., 2004), DWQ (21; Freeman et al., 2020), and the summed LSAS Avoidance and Endorsement subscales (30, mild social anxiety; Liebowitz, 1987). For each participant and caregiver, we determined whether the total measure score met or exceeded the designated clinical threshold and assessed whether the pair agreed. For each measure, we fit a logistic regression on the binary outcome of participant-caregiver agreement or disagreement relative to the clinical threshold, with IQ and age as predictors (**Model 3**).

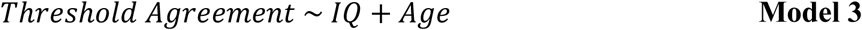

## 3 Results

### 3.1 Between-Measure Correlations

In both groups, we observed moderate (R=0.3-0.49) to strong (R≥0.50) correlations between most measures (**Figure 2**). This reflects general population patterns (Freeman et al., 2012), supporting the validity of both self and caregiver-report. Although we did not formally test pairwise differences in correlation strength between groups, some qualitative differences in the direction of correlations were apparent (**Figure 2C**). The LSNS-6 total score, reflecting the robustness of social networks, showed the greatest divergence. In both groups, more robust social networks were strongly negatively correlated with social avoidance (LSAS Avoidance), and moderately negatively correlated with both social anxiety (LSAS Fear) and social cognition problems (DACOBS-SOC). In the caregiver group, social network robustness was also moderately negatively correlated with most other measures (**Figure 2A**). However, in the WS group, social network robustness was positively correlated with these measures (**Figure 2B**). These results suggest that associations between social networks and measures of paranoia, delusional thought content, perseveration, worry, emotion regulation and cognitive bias may differ depending on whether the report comes from the individual or the caregiver - an important consideration, given that atypical social insight and motivation are core features of the WS phenotype.

**Figure 2.**
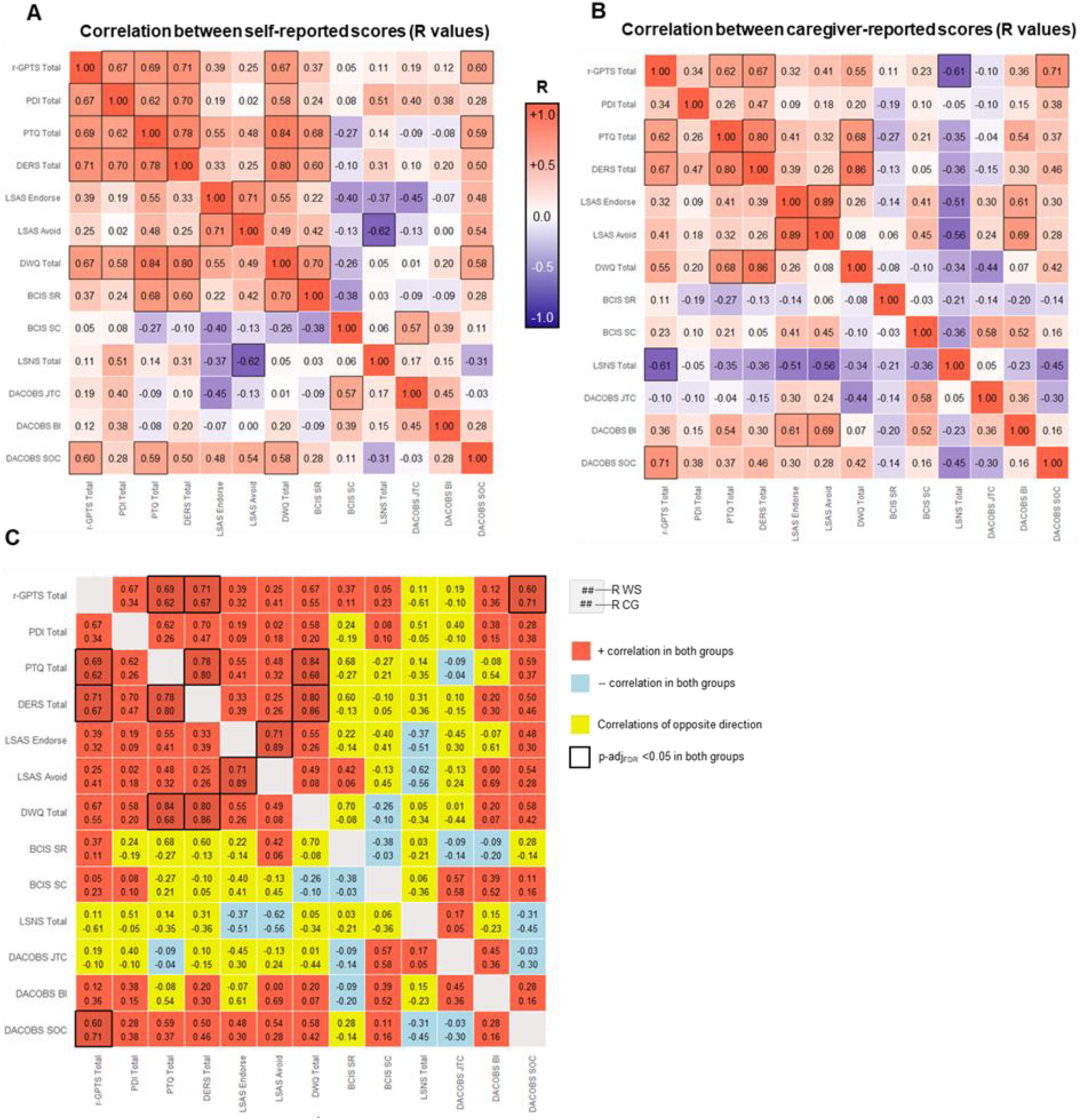
Between-Measure Correlations in Self- and Caregiver-Report. **A.** Pairwise correlations (Pearsons’s correlation coefficient, R) between measures in the participant (WS) sample. **B.** Pairwise correlations between measures in the caregiver sample. In both heatmaps, redder colors represent a stronger positive correlation (larger positive R value), whiter colors represent no correlation (R value near zero), and bluer colors represent a stronger negative correlation (larger negative R value). Statistically significant correlations (p-adj_FDR_<0.05) are outlined in black. **C.** Correlations for both groups on a single heatmap, with colors demonstrating whether correlations were in the same direction in both groups (positive = red, negative = blue) or in opposite directions across groups (yellow). Statistically significant correlations (p-adj_FDR_<0.05) in both groups are outlined in black.

### 3.2 Participant-Caregiver Total Score Agreement

The mean and standard deviation of self- and caregiver-reported scores for all measures are reported in **Table 2**. Paired Wilcoxon sign rank tests revealed no significant participant-caregiver differences in total scores (**Table 2, Supplementary Figure 1**), except for delusional ideation as measured by the PDI-21 (W=136, FDR-adjusted p-value=0.008). Notably, participants scored significantly higher on the endorsement of delusional ideation compared to their caregivers (**Figure 3A**), suggesting that caregivers may not be fully aware of the adult with WS’s unusual beliefs. Participant-caregiver agreement and disagreement across the 21 beliefs assessed in the PDI is shown in **Figure 3B**. Interestingly, some of the most unusual and bizarre beliefs assessed on the scale were endorsed only by participants, suggesting caregivers are unaware of these experiences (e.g., “Have your thoughts ever been so vivid that you were worried other people could hear them?”).

**Figure 3.**
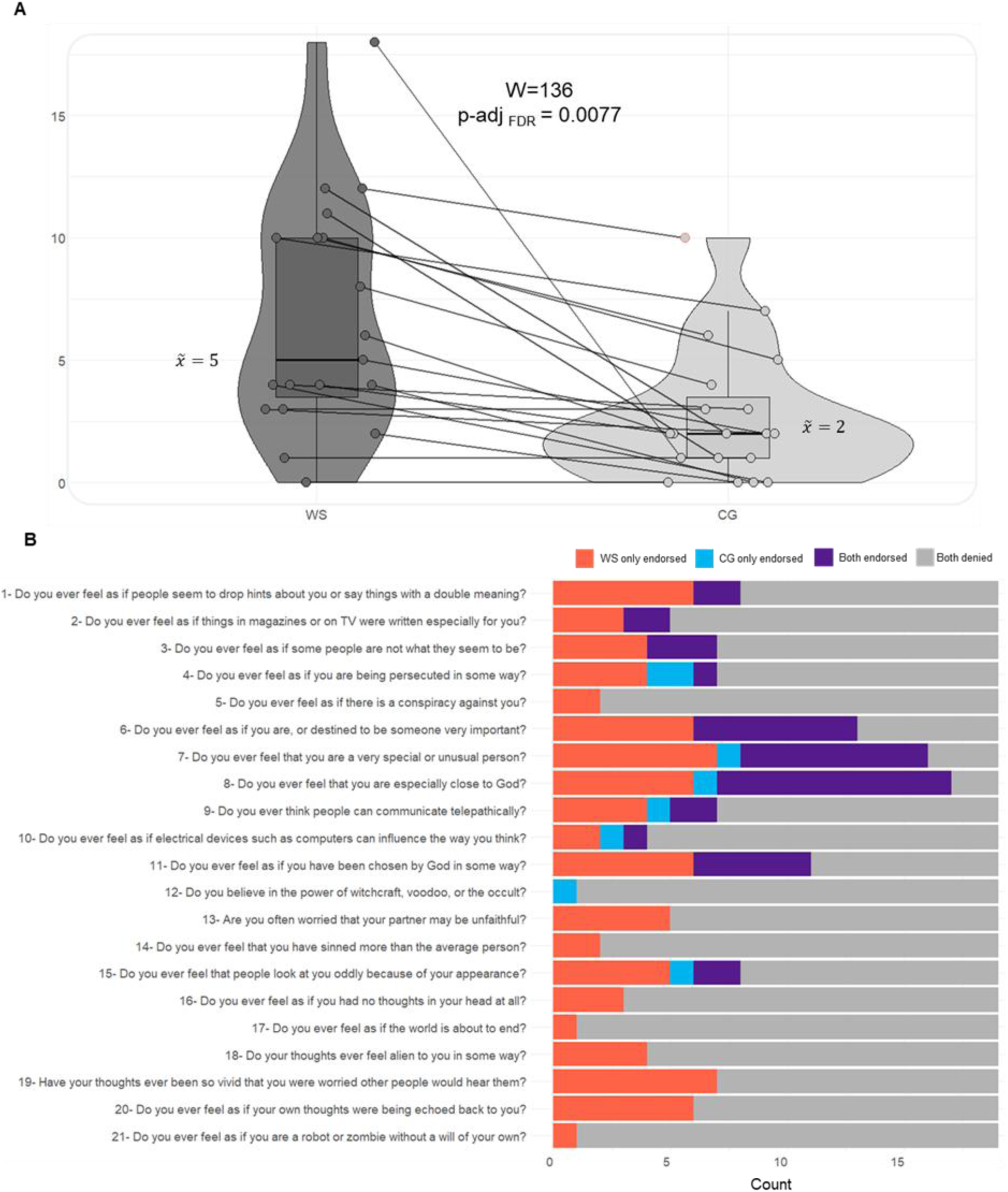
PDI-21 Endorsement in WS and Caregivers. **A.** Pairwise relationships across groups; all participants endorsed at least the same or greater number of unusual beliefs was reported by their caregiver. **B.** Count of participant-caregiver pairs in each of the following categories for every belief. Red: belief only endorsed by participant (WS); blue: belief only endorsed by caregiver (CG); purple: belief mutually endorsed; gray: belief mutually denied.

**Table 2.**
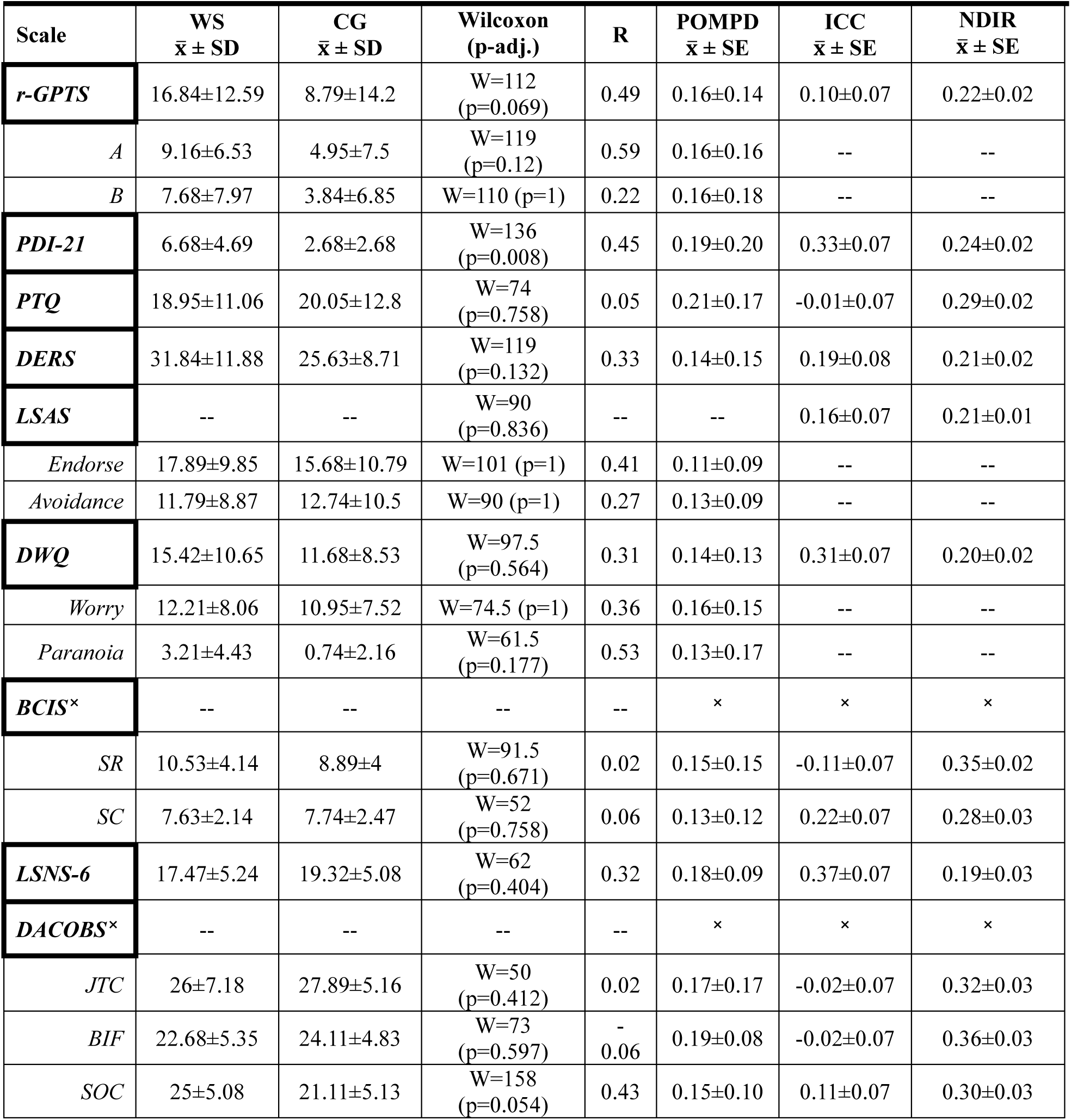
Statistics by Measure. The statistics computed for each measure (total score and/or subscale score) are shown. This includes: group averages for participants (WS) and caregivers (CG), results of the paired Wilcoxon signed-rank tests comparing differences in participant-caregiver total scores, Pearson’s correlation coefficient (R) between participant-caregiver total scores, the average percent of maximum possible difference (POMD) between participant-caregiver total scores, and the average interclass correlation (ICC) between participant-caregiver responses at the level of individual items and the average normalized differences in item-level responses (NDIR).

When examining correlation (R) in total score between participants and caregivers (**Table 2**, **Figure 4**), we found moderate-to-strong correlations (range 0.221-0.595) across all measures, except for those of perseveration (PTQ; R=0.05), cognitive insight (BCIS-SC; R=-0.061; BCIS-SR; R=-0.017), and the cognitive biases of jumping-to-conclusions (DACOBS JTC; R=0.0254) and belief inflexibility (DACOBS BIF; R=-0.0611).

**Figure 4.**
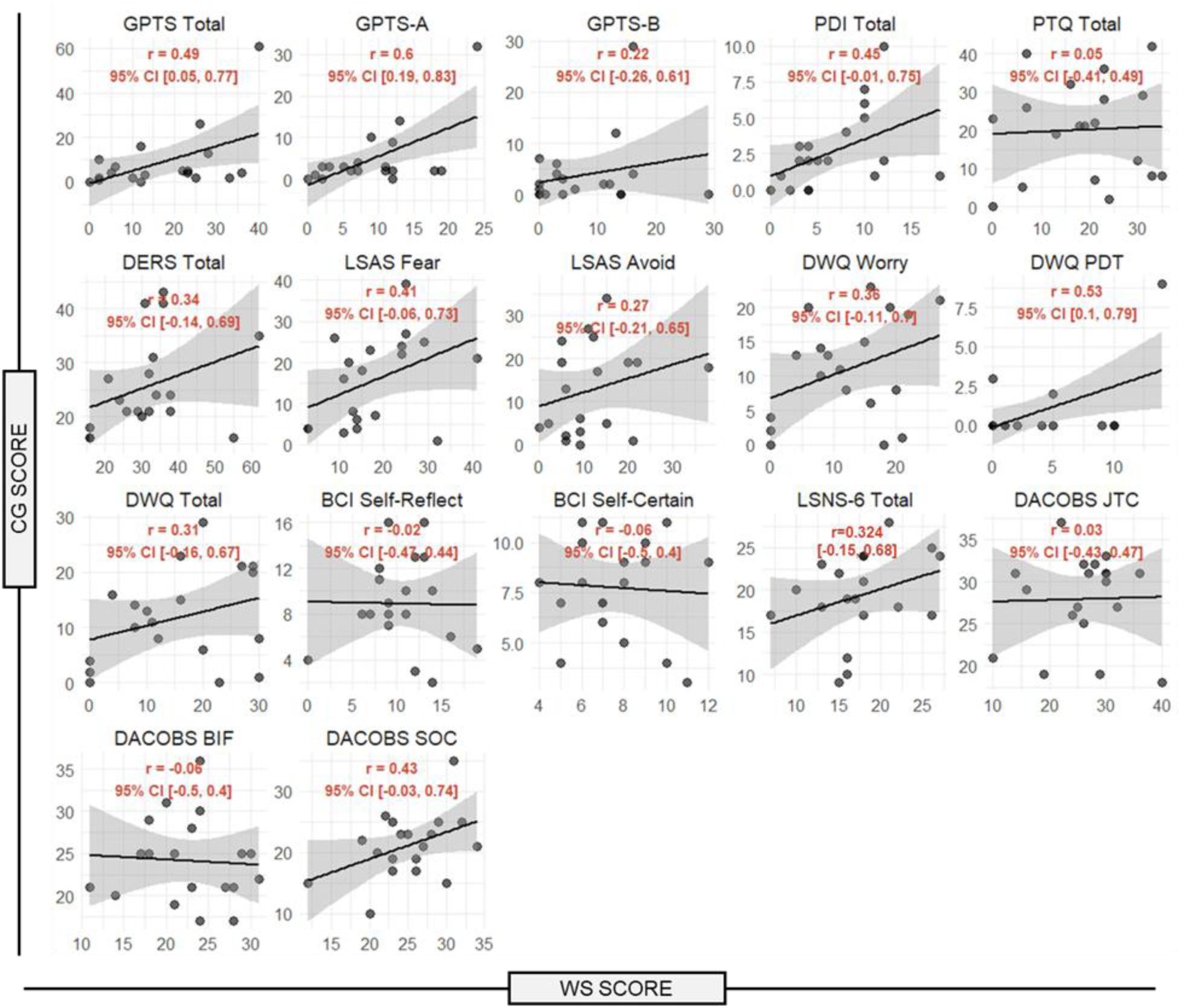
Self- and Caregiver-Report Total Score Correlations. Pearson’s correlation coefficient (R) between the total scores for the participant (self-report, horizontal axis) and caregiver (caregiver-report, vertical axis) for all measures and subscales.

Regarding POMPD, our mixed effects model (Model 2) revealed no significant main effect of measure nor IQ (B=-0.0015, SE=0.0009, p=0.1304) on participant-caregiver agreement (**Supplementary Table 4**, **Figure 5**). However, contrary to our hypothesis, there was a main effect of age (B=0.005, SE=0.0022, p=0.0348) such that older age was associated with worse agreement (greater POMPD). As a *post hoc* exploratory analysis, we specifically conducted a general linear model examining the effect of age and IQ on PDI-21 POMPD values, given that this was the only measure found to significantly differ across groups on the paired Wilcoxon sign rank test. As with the primary mixed effects model, the *post hoc* analysis revealed no significant effects of IQ (B=-0.0003, SE=0.0026, p=0.8890), but a significant effect of age (B=0.0166, SE=0.0060, p=0.0140) on the POMPD in PDI-21 total score.

**Figure 5.**
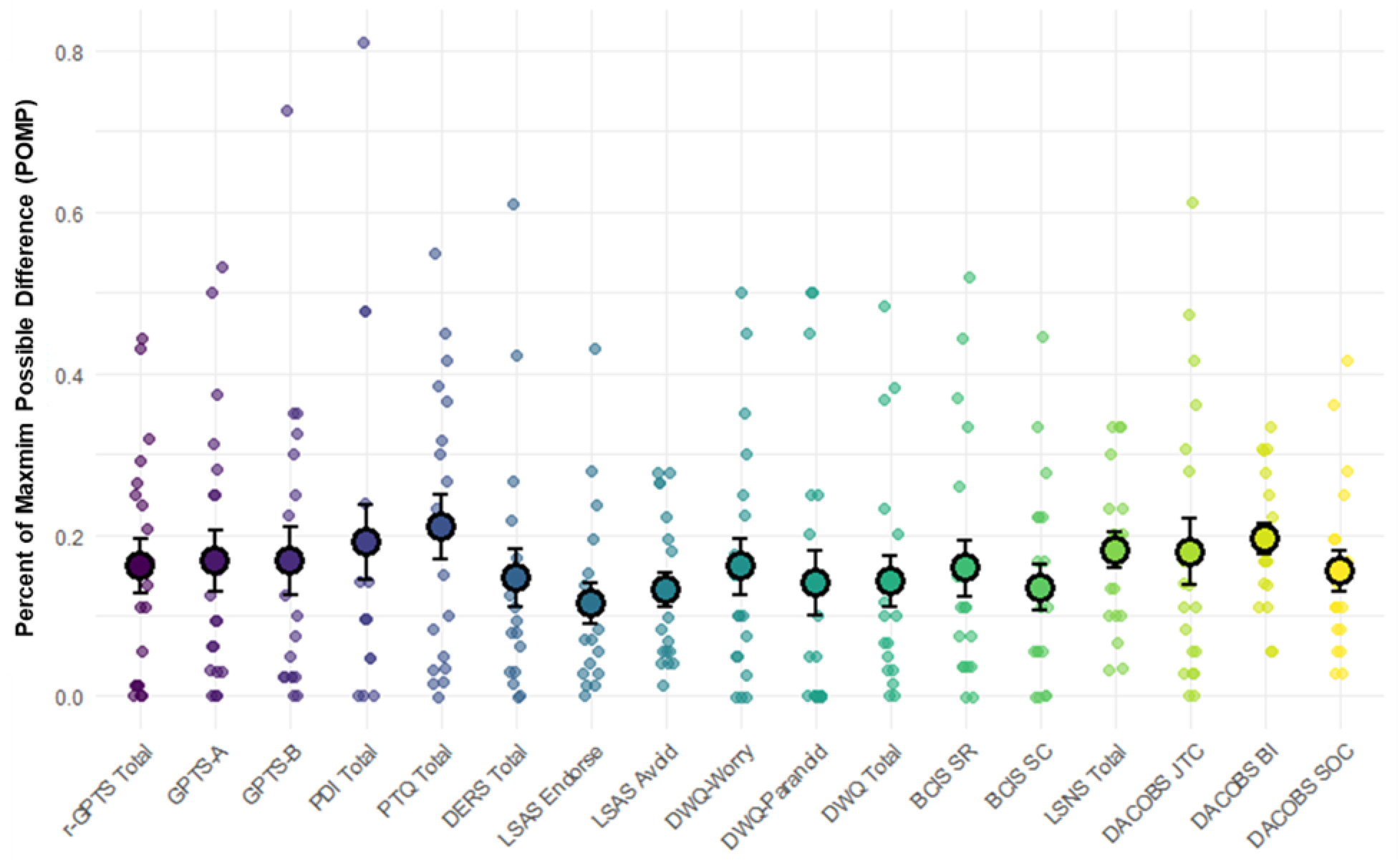
Percent of Maximum Possible Difference Scores. Mean and standard deviation of the percent of maximum possible difference (POMPD) for total and subscale scores.

### 3.3 Participant-Caregiver Item-Level Agreement

Although participant-caregiver differences in *total* scores were non-significant (except the PDI-21), we were also interested in the degree of agreement on an item-by-item level. Across measures, item-level ICCs were generally poor (<0.5, Koo & Li, 2016; **Table 2**, **Figure 6A**). NDIRs were not influenced by age (B=0.0059, SE=0.0027, p=0.0509) or IQ (B=-0.0023, SE=0.0013, p=0.0940). However, they differed significantly between measures. To formally test these differences, we used the *emmeans* package to compute estimated marginal means of NDIR across measures (**Figure 6B**) and conducted pairwise comparisons with FDR correction. Several statistically significant differences across measures emerged (**Figure 6C**). Specifically, the PTQ (emm=0.29), BCIS-SR (emm=0.35), BCIS-SC (emm=0.28), and DACOBS subscales (JTC emm=0.32; BIF emm=0.36; SOC emm=0.30) had significantly greater mean NDIR compared to most other measures, reflecting worse participant-caregiver agreement at the item-level. In contrast, the DERS (emm=0.21), LSAS (emm=0.20), DWQ (emm=0.20), and LSNS (emm=0.19) had significantly lower mean NDIR compared to many other measures, reflecting better item-level agreement.

**Figure 6.**
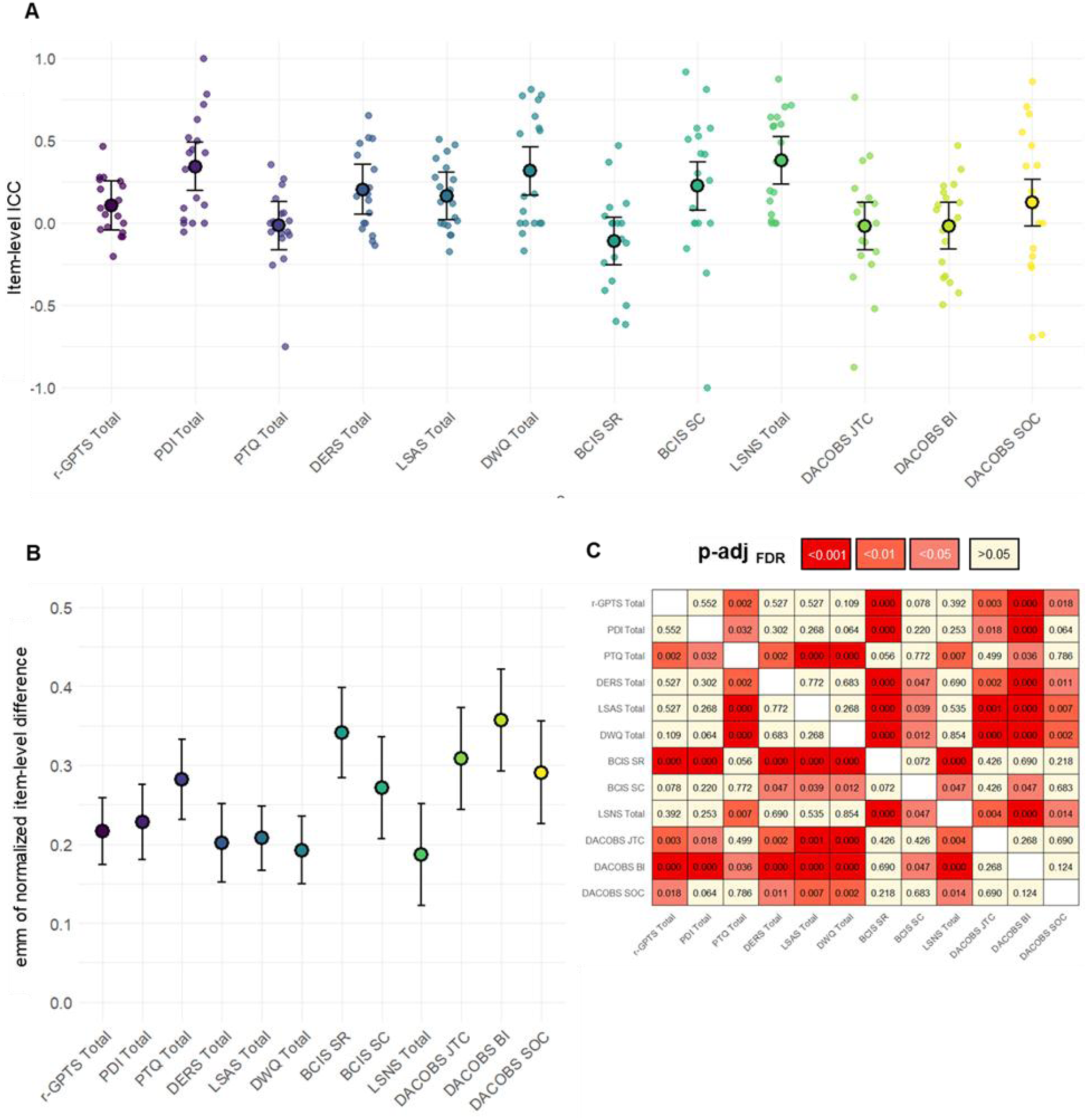
Item-Level Agreement. **A.** Item-level ICCs across participant-caregiver pairs for each measure. **B.** Estimated marginal mean of the normalized differences in item-level responses (NDIR) across measures. Note that higher NDIR values reflect worse participant-caregiver agreement. **C.** Heatmap of pairwise comparisons (p-values) of NDIR scores across measures. The PTQ, BCIS subscales, and DACOBS subscales demonstrate higher NDIR values than multiple other measures, indicating that these measures have the poorest item-level agreement.

The key outcomes from this analysis are twofold. First, the overall poor item-level ICCs demonstrate that although total scores may be similar between participants and their caregivers, the responses to individual items composing the total scores may not necessarily be similar. This may be of particular importance for measures such as the PDI-21 or DACOBS, in which different items represent distinct delusions or biases, and in which differences in endorsed items may be important. Second, as hypothesized, the measures with the lowest inter-rater reliability concern cognitive insight, cognitive bias, and perseveration, which are largely internal processes. In contrast, emotion regulation, anxiety, worry, and social networks—domains which may be more likely to result in externalized behavior—demonstrate higher inter-rater reliability.

### 3.4 Clinical Thresholds

Our final analysis concerned the likelihood of reaching a clinical threshold, and whether this differed between participants and their caregivers. On the r-GPTS, 37% (n=7) of participant-caregiver pairs disagreed on threshold outcome; in all seven cases, self-report exceeded the clinical threshold while caregiver-report did not (**Figure 7A**). On the PDI-21, 32% (n=6) of pairs disagreed; again, in all six cases, self-report exceeded the threshold while caregiver-report did not (**Figure 7B**). On the DWQ, outcomes were more variable: 37% (n=7) of pairs disagreed, but there were five instances of self-report exceeding the clinical threshold and two instances of caregiver-report exceeding threshold (**Figure 7C**). On the LSAS, 26% (n=5) of pairs disagreed, including four instances in which caregiver-report exceeded threshold and one instance in which self-report exceeded threshold (**Figure 7D**).

**Figure 7.**
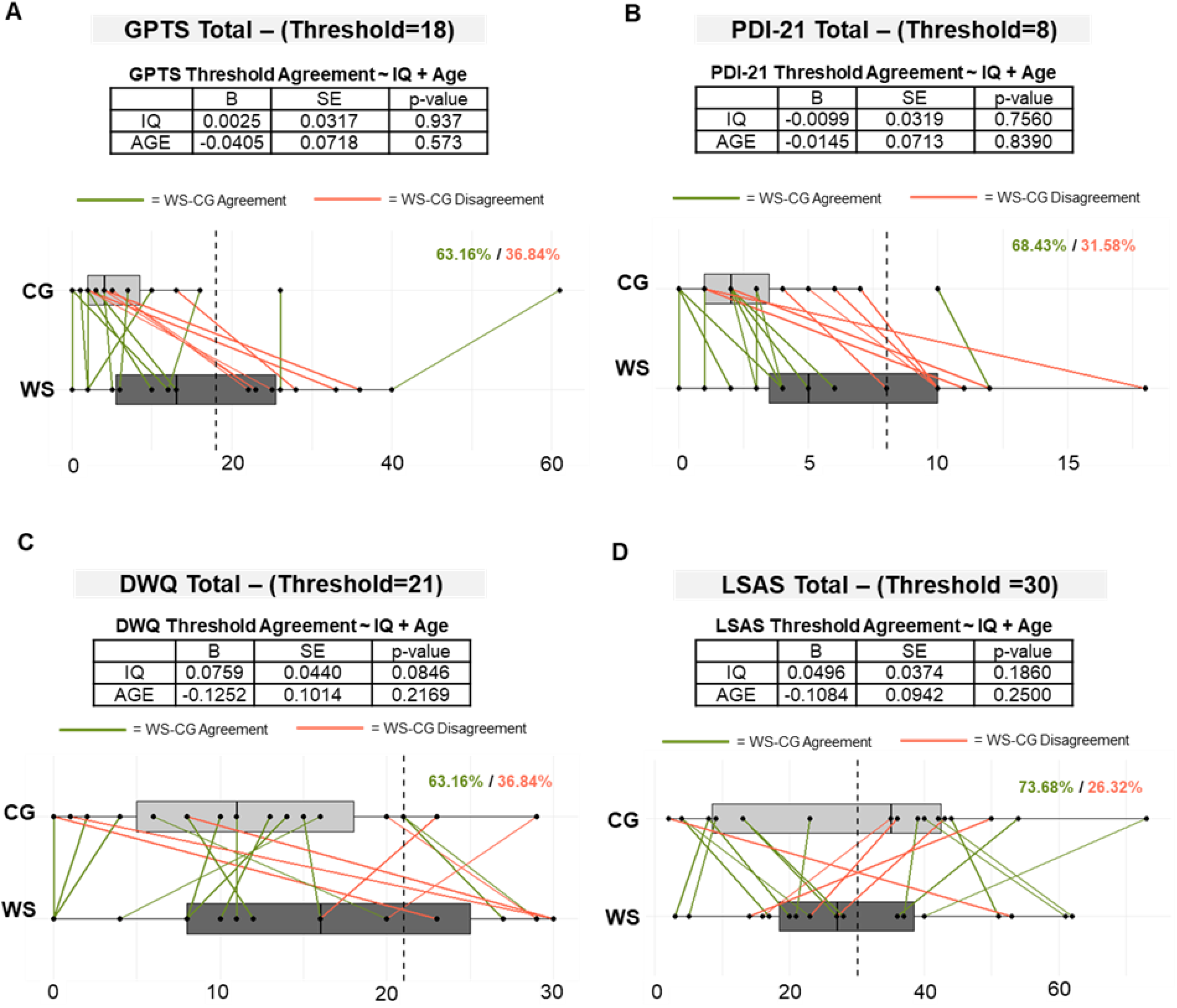
Clinical Threshold Agreement. Paired box plots show participant and caregiver total scores on **A.** r-GPTS, **B.** PDI-21, **C.** DWQ, and **D.** LSAS. For each pair plot, participant-caregiver dyads who agreed on threshold outcome are plotted in green; those who disagreed are plotted in red. For each clinical assessment shown here, contrast estimate (β), standard error (SE), and p-value from each logistic regression are shown for the predictor variables of interest, IQ and age.

As shown in **Figure 7**, age and IQ did not predict participant-caregiver clinical threshold agreement for any measure. These results suggest that even while paired comparisons of scores across groups revealed no differences, there may be discrepancy between adults with WS and their caregivers in reporting relevant to clinically meaningful thresholds. Failure to collect self-report may result in lack of or underreporting of clinically significant paranoia and delusional ideation, as well as discrepant reporting of worry and anxiety.

## 4 Discussion

In this study, we characterized and compared self- and caregiver-reported experiences of thought disturbances, as well as associated cognitive-affective risk factors, in 19 adults with WS and mild-to-moderate intellectual disability. We observed that both self- and caregiver-report revealed clinically significant paranoia, delusions, worry, and social anxiety in several individuals, suggesting that at least a subset of adults with WS and intellectual disability exhibit a profile of thought disturbances coincident with associated risk factors. To the authors’ knowledge, this is the first study to systematically examine these traits in the WS population.

Both self- and caregiver-report showed strong inter-correlations among measures consistent with previously established patterns, supporting the validity of both perspectives in reporting thought disturbances and associated factors. While paired Wilcoxon signed-rank tests revealed no systematic differences between participant- and caregiver-reported total scores (except on the PDI-21), the amount of correlation between participants and caregiver total scores varied across measures, ranging from strong to near zero. Item-level analyses (ICCs and NDIRs) also indicated moderate-to-poor reliability, with the lowest agreement for measures of perseveration (PTQ), cognitive insight (BCIS), and cognitive bias (DACOBS). These results suggest that although participant and caregiver reports align in estimating overall symptom burden, they often diverge at the level of individual responses. Thus, the value of collecting both self- and caregiver-reports may depend on the instrument and whether item-level detail is needed for clinical decision-making. Importantly, discrepancies in agreement were not explained by participant age or IQ. Finally, analysis of clinical thresholds showed that participants were more likely than caregivers to endorse clinically significant paranoia and delusional ideation, raising concerns that reliance on caregiver report alone may underestimate risk and lead to missed opportunities for diagnosis and intervention.

### 4.1 The value of collecting self-report in individuals with Williams Syndrome

Taken together, these findings suggest that self-report of paranoia, delusions, and related cognitive-affective processes (e.g., rumination, emotion regulation, anxiety, worry, cognitive insight, cognitive bias) is valid in WS, and generally converges with caregiver report at the level of total scores. However, it remains unclear as to whether the added time and resources required to adapt self-report tools in this population yield additional clinical benefit. Our analysis of item-level agreement offers important insight in this regard. ICCs demonstrated that while total scores may appear comparable, agreement on individual items was generally poor to moderate, and varied significantly by measure. As expected, when comparing item-level agreement across measures, those with the worst agreement generally assessed internal cognitive experiences (perseveration, cognitive insight, and cognitive bias), while those with the best agreement assessed more externalized behaviors (emotion dysregulation, anxiety and avoidance, worry, and social networks). The finding of no significant differences in total scores juxtaposed with low item-level ICC suggests that while participant and caregiver report are generally reliable in terms of the overall severity or magnitude of assessed symptoms, they are not necessarily matched at the level of individual items. Thus, for instruments in which the specific nature of endorsed items is important for informing clinical risk assessment or treatment planning it may be best-practice to collect both self- and caregiver-report.

### 4.2 Spotlight on delusional ideation

Prior to this study, the authors were unaware of any research explicitly examining the presence of thought disturbances in WS, despite the presence of other cognitive features known to be related to the incidence of paranoia and delusional ideation. Our results suggest the degree of delusional ideation in this population may be more significant than previously believed. Internal experiences such as thought disturbances are particularly challenging for proxies to assess, as accuracy depends on both insight into the individual’s internal narrative and the quality of communication, both of which may be limited for individuals with neurodevelopmental disorders and intellectual disabilities. In our sample, participant and caregiver total scores largely agreed on all measures except the PDI-21, where participants endorsed significantly more delusional ideation than their caregivers. Whether this reflects overreporting of unusual beliefs by participants (e.g., due to acquiescence response bias; (Emerson et al., 2013)) or underestimation by caregivers is unclear; however, given that PDI-21 scores correlated with other measures as expected, we interpret participant self-report of delusional ideation as both valid and reliable.

Beyond disagreement at the level of total scores, the PDI-21 also warrants special attention at the item-level. While the ICC of item-level responses for the PDI-21 was acceptable (0.33), this should be cautiously interpreted given the significant pairwise differences in total scores. The PDI-21 endorsement items are scored either 0 or 1; therefore, for every item, there is a 50% probability that individuals with WS will agree with their caregiver. This is much higher than for a 4- to 7-point Likert scale, as is the case for the remaining measures, leading to relative inflation of the PDI-21 ICC. As just discussed, for instruments in which the nature of the individual items is important—such as the distinct types of unusual beliefs on the PDI-21—failure to collect self-report may lead to clinical misunderstanding of the unusual beliefs. These results therefore suggest that for assessing both the severity and the nature of delusional ideation in individuals with neurodevelopmental disorders and intellectual disabilities, the additional effort to collect self-report of unusual beliefs is necessary to accurately characterize both the severity and content of these beliefs.

### 4.3 Optimizing accessibility of self-report methodology in individuals with neurodevelopmental disorders and intellectual disabilities

Given the generally low item-level reliability and significant differences in normalized item-level reliability across measures, there is an outstanding question as to what drives these differences. Notably, measures with the poorest agreement (BCIS, DACOBS) rely on agreement-based Likert scales (e.g., “Strongly agree” to “Strongly disagree”), which often involve negations or complex phrasing. For individuals with intellectual disabilities, such formats may be particularly confusing. For example, negative clauses such as, *“I do not need to consider alternatives when making a decision,”* requires reversing the clause to answer accurately. In contrast, scales that assess frequency (e.g., “Almost never” to “Almost always”) showed higher inter-rater reliability (DERS, LSAS, DWQ), likely because they are more concrete and easier to interpret (Hartley and MacLean Jr., 2006).

These findings underscore the need for adapting self-report measures for individuals with neurodevelopmental disorders and intellectual disabilities, as well as carefully considering how the structure of response scales, instrument length, and language may impact the quality and integrity of response. Previous work shows that simplifying language (Vlot-van Anrooij et al., 2018), adding visual supports, and allowing verbal response formats can improve comprehension and reliability (Walton et al., 2022). For example, a recent adaption of the self-reported Adaptive Behavior Assessment System, 3^rd^ Ed. (“Adaptive Behavior Assessment System: Third Edition | SpringerLink,” n.d.) demonstrated that simplified prompts and visual aids enhanced participant understanding, recall, and reliability with caregiver report across all domains (Kooijmans et al., 2024). In line with this, we show that with appropriate adaptation, self-reports of thought disturbances can be successfully and reliably administered in individuals with WS. Importantly, beyond highlighting specific adaptations which may be useful for individuals with intellectual disabilities, this work points towards the need to broadly consider the accessibility of self-report measures for use in any population with cognitive limitations.

### 4.4 Limitations and Future Directions

Although our sample size (*N*=19) was large relative to other work in the WS population, to better understand inter-rater reliability in this population, an effort should be made to recruit a significantly larger sample as well as a wide variety of ages, the latter of which would allow researchers to understand how developmental trajectory affects item-level and total score discrepancies. Similarly, while we provide a preliminary socio-cognitive profile of thought disturbances and associated risk factors in adults with WS and ID, a larger sample is required to generate more accurate estimates of population-level rates of these traits as well as to assess the impact of demographic covariates such as age and sex. We did not confirm a genetic diagnosis of WS through medical record review; diagnosis was confirmed through caregiver report. Participants varied in daily proximity to caregiver - some participants lived with their caregivers, while others lived independently or in supportive living facilities, differences which may have impacted the degree of agreement within pairs. Finally, while there is a strong consensus that individuals with WS demonstrate heightened generalized anxiety and phobias relative to the general population (Dykens, 2003; Woodruff-Borden et al., 2010), reports on social anxiety in WS are mixed (Binelli et al., 2014; Riby et al., 2014; Royston et al., 2021). In the present study, we focus on social anxiety rather than generalized anxiety because social anxiety has been linked more specifically to thought disturbances such as paranoia and delusional ideation (Schutters et al., 2012; Tone et al., 2011). Nonetheless, it will be important for future work in this population to assess additional anxiety domains, given their consistency with the broader WS phenotype.

Despite these limitations, to our knowledge this is the first study to demonstrate the validity of self-reported thought disturbances in adults with WS and ID. These findings lay important groundwork for future investigations into the unique psychiatric profile of this population, and highlight the need to examine how thought disturbances may interact with cognitive, social, and functional outcomes in adults with WS.

## 5 Conclusions

This study provides preliminary evidence of elevated paranoia, delusions, and atypical patterns of social cognition, perseveration, belief inflexibility, and cognitive insight in adults with WS and intellectual disabilities. Our findings also demonstrate that self-report is a valid and valuable method for assessing these psychopathologies. While caregiver- and self-report converge on overall severity of cognitive disturbances, caregiver-report alone may fail to capture clinically significant symptoms of paranoia and unusual beliefs. In particular, caregivers tend to under-report delusional ideation, which may contribute to under-detection of this thought disturbance in WS, obscure related challenges in interpersonal and occupational functioning, and lead to missed opportunities for accurate diagnosis and timely intervention. Although collecting both self- and caregiver-report is resource intensive, it provides a more comprehensive clinical picture and supports more accurate characterization of the unique cognitive and psychiatric profile of individuals with WS. Importantly, the validity and accuracy of self-report in individuals with neurodevelopmental disorders and intellectual disabilities can be strengthened using adapted measures and administration methods designed to enhance accessibility. These principles should be considered not just in the context of neurodevelopmental disorders and intellectual disabilities, but broadly in the use of self-report with any population with impaired cognitive function.

## Data Availability

The datasets generated and analyzed during the current study are available from the corresponding author on reasonable request.

