## Supplementary File for "Agreement Between Self- and Caregiver-Report of Thought Disturbances in Adults with Williams Syndrome and Intellectual Disability"

**Supplementary Figure 1. Paired Distributions of All Measures**
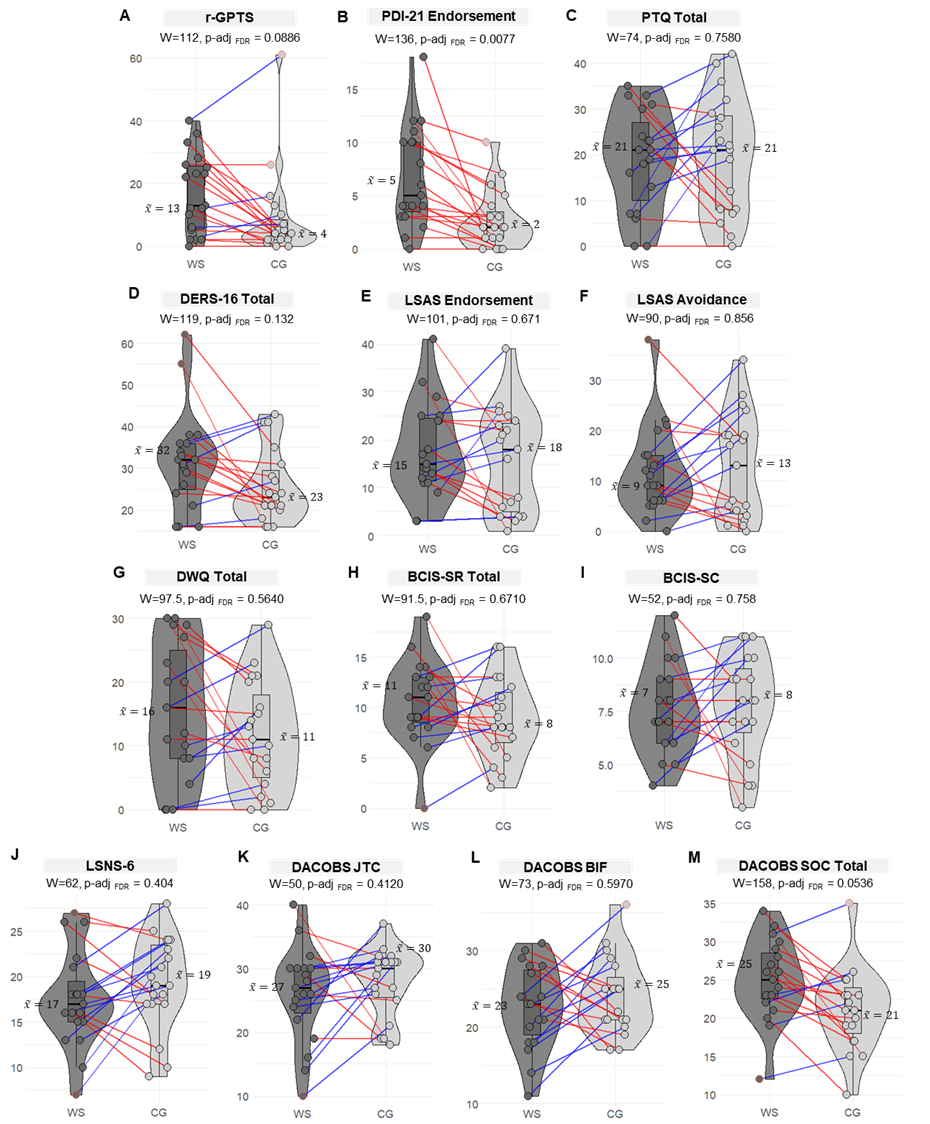

**Supplementary Figure 1.** Pair plots illustrating the self- and caregiver-reported scores for all measures of interest. **A.** r-GPTS total score. **B.** PDI-21 Endorsement score. **C.** PTQ total score. **D.** DERS-16 total score. **E.** LSAS Endorsement score. **F.** LSAS Avoidance score. **G.** DWQ total score. **H.** BCIS Self-Reflectiveness (SR) score. **I.** BCIS Self-Certainty (SC) score. **J.** LSNS-6 total score. **K.** DACOBS Jumping to Conclusions (JTC) score. **L.** DACOBS Belief Inflexibility (BIF) score. **M.** DACOBS Social Cognition Problems (SOC) score.

**Supplementary Table 1: Assessment Battery**

| **Measure** | **Domain Assessed** | **Completed By** |
| --- | --- | --- |
| *Green Paranoid Thought Scale (GPTS)* | Paranoia | Participant and Caregiver |
| *Peters et al. Delusion Inventory-21 (PDI-21)* | Delusions | Participant and Caregiver |
| *Perseverative Thinking Questionnaire (PTQ)* | Perseveration, Rumination | Participant and Caregiver |
| *Difficulty in Emotion Regulation Scale (DERS)* | Emotion Regulation | Participant and Caregiver |
| *Liebowitz Social Anxiety Scale (LSAS)* | Social Anxiety | Participant and Caregiver |
| *Dunn Worry Questionnaire (DWQ)* | Worry | Participant and Caregiver |
| *Beck Cognitive Insight Scale (BCIS)* | Cognitive Insight | Participant and Caregiver |
| *Lubben Social Network Scale (LSNS-6)* | Socialization | Participant and Caregiver |
| *Davos Assessment of Cognitive Biases Scale (DACOBS)* | Cognitive Biases | Participant and Caregiver |
| *Kauffman Brief Intelligence Test-2 (KBIT-2)* | Intelligence | Participant only |
| *Vineland Adaptive Behavior Scale (VABS-3)* | Adaptive Functioning | Caregiver only |
| *Demographic Survey* | Demographics | Caregiver only |
| *Psychiatric History* | Clinical History | Caregiver only |
| **Supplementary Table 1.** Each assessment administered is shown below, including the abbreviated name, the cognitive-affective domain assessed, and who completed the assessment (participant and/or caregiver). | | |

**Supplementary Table 2 –** All self-reports were administered to participants by trained proctors (KB, EL). Each question and response item was read aloud. Proctors were attentive to cues suggesting comprehension challenges, including lack of or extensive response delay, nonverbal signs of frustration (e.g. brow furrowing, grimacing), or discrepant answers across items assessing the same construct. Below, we provide a list of the general domains of adaptations, followed by specific example of these adaptations. This is not an exhaustive nor comprehensive list of all adaptations made, but rather an illustration of the nature of these adaptations such that they can be flexibility replicated.

**General domains of adaptation**

1. **Simplification of vocabulary**: Defining or strategically substituting words reflecting advanced vocabulary or abstract concepts (e.g. “gossiping”, “critically”, “conspiracy”, “persecuted”, “destiny”, “hostile”).
2. **Providing illustrative examples:** Providing an example of a situation or context which illustrates the impetus of the question.
3. **Asking follow-up to confirm comprehension:** Asking follow-up questions to confirm understanding of the item content; this might include asking the participant to provide an example to confirm comprehension.
4. **Restructuring a double negative:** If providing a response requires a double negative, the wording of the question may be restructured for clarity.
5. **Adapting experiences which might be unrelatable:** For items which involve scenarios or examples, adapting the prompt if it is not relevant to the participant’s daily activities.
6. **Rewording:** As a last resort, there are rare items in which the wording was more dramatically restructured to improve clarity.

| **Domain** | **Measure** | **Original Item** | **Adaptation** |
| --- | --- | --- | --- |
| 1 | *r-GPTS* | People have been hostile towards me on purpose. | Last week people were rude, mean, or angry to me on purpose” |
| 1 | *PDI-21* | Do you ever feel as if you are being persecuted in some way? | Do you ever feel as if someone is trying to hurt you or harm you because of who you are, or what you believe? |
| 3 | *PDI-21* | Do you ever think people can communicate telepathically? | Who can communicate telepathically? Have you ever done it? With whom? |
| 2 | *PTQ* | My thoughts prevent me from focusing on other things. | My thoughts prevent me from focusing on other things. Sometimes when I am at work, at school, or doing chores, I can’t get things done because of my thoughts. |
| 4 | *PTQ* | My thoughts are not much help to me. | My thoughts help me. How often do your thoughts help you – never, rarely, sometimes, often, or almost always? |
| 2 | *PTQ* | When I am upset, I have difficulty controlling my behaviors. | When I am upset, I have difficulty controlling my behaviors. I can’t stop yelling, crying, hitting or running away. |
| 2 | *LSAS* | Calling someone on the phone while you are in public - speaking on the phone in a public place | Let’s pretend you are at the grocery store and you have to call your Mom (or Dad) to ask about buying apples. How scared or afraid would that make you feel? |
| 5 | *LSAS* | Writing while being observed - for example, signing paperwork | Writing while being observed – for example, signing paperwork or forms, or signing a birthday card while someone is watching you so they can sign it next. |
| 2, 5 | *LSAS* | Returning goods to a store where returns are normally accepted | Let’s pretend you bought a shirt that did not fit and you had to return it to the store. How scared would that make you feel? Would you avoid going to the store and keep the shirt even though it did not fit? |
| 1 | *DWQ* | It has been hard to clear my head of suspicions. | It has been hard to clear my head of thoughts that other people are up to no good. |
| 8 | *DWQ* | Anything and everything has set my mind thinking about people trying to upset me. | People are trying to make me sad or angry, and I think about it a lot. |
| 1, 2 | *BCIS* | Some of my experiences that have seemed very real may have been due to my imagination. | Some of my experiences that have seemed very real may have been due to my imagination. Some things that happen might be all in my head. |
| 3 | *BCIS* | Other people can understand the cause of my unusual experiences better than I can. | Who can understand your experiences better than you can? |
| 4 | *DACOBS* | I don't need long to reach a conclusion. | I can make a choice quickly. |
| 1, 2 | *DACOBS* | The right conclusion often pops in my mind | The right conclusion often pops in my mind. The right answer pops into my head like a lightbulb! |
| 4 | *DACOBS* | I don't need to consider alternatives when making a decision. | I want to look at all the options before I make a choice. |
| 8 | *DACOBS* | I avoid considering information which will disconfirm my beliefs. | I don’t want to listen to any information that goes against what I think or what I believe. |

**In addition to these adaptations to the measures, two specific adaptations were also made to the order of item administration, when necessary:**

- **Skip & return:** If there was significant difficulty achieving comprehension of an item, or the participant cannot decide on a response, the item was skipped and returned to at the end of the assessment. Responses were not entered until comprehension was displayed through clarifying questions.

**Revisiting prior responses:** Many items within measures assess similar ideas or constructs. If directly opposing responses are given, prior responses were revisited and reexplained to clarify that understanding was achieved.

**Supplemental Table 3: Norms, clinical means, and internal consistency of self-report measures**

|  | **Normative Samples** | **Clinical** $\bar{\boldsymbol{x}}$ | **Internal Consistency (Cronbach’s α)** |
| --- | --- | --- | --- |
| *r-GPTS^1^* | N=353 HC,  N=50 with persecutory delusions | r-GPTS-A: 46.4±16.4  r-GPTS-B: 55.4±15.7 | 0.90 |
|  |  |  | 0.90 |
| *PDI-21^2^** | N=444 HC,  N=33 with hx of delusions | 33.9±8.4 | 0.82 |
| *DWQ^3^* | N=250 HC,  N=50 with hx of persecutory delusions** | General: 27.3±7.48  Paranoid: 12.7±5.05 | 0.93 |
| *LSAS^4^* | N=382 with social phobia*** | Fear: 35.5±13.6  Avoidance: 31.6±14.5 | 0.95 |
| *PTQ^5^* | N= 501 HC,  N=113 with various untreated psychiatric diagnoses (original German validation) | Not provided | 0.95 |
| *DERS-16^6^* | N=357 HC | -- | 0.93 |
| *BCIS^7^* | N=150 with schizophrenia, schizoaffective disorder, or major depressive disorder with and without psychosis | SR: 14.01±4.84  SC: 6.99±3.5 | SR: 0.68  SC: 0.60 |
| *LSNS-6^8^* | N=7,432 HC seniors across 3 sites | -- | 0.70-0.83 |
| *DACOBS^9^*  ****** | N=186 HC,  N=138 (sample 1) and N=71 (sample 2) with schizophrenia spectrum disorders (SDD) | JTC: 25.59±6.6  BIF: 20.72±6.82  SOC: 24.28±7.02 | JTC: 0.72  BIF: 0.74  SOC: 0.76 |
| **The PDI-21 also assesses the domains of distress, preoccupation, and conviction for each endorsed item. In this study, we utilize only data from the endorsement subscale; however, the normative information provided here reflects the full measure*  ***Cross validation in 273 HC, 79 individuals with persecutory delusions, 93 individuals with social anxiety*  ****Original 1987 publication of LSAS did not include normative data. First study providing normative data is reported here and in* ^10^  *****Full DACOBS scale includes 7 subscales, each comprising 6 items; only 3 subscales were included in this analysis (Jumping to Conclusions – JTC; Belief Inflexibility – BIF; Social Cognition Problems - SOC)* | | | |
| **Supplemental Table 3:** For each measure, the population in which the measure was normed is described including the number of healthy controls (HC) and number of individuals with a clinical diagnosis or history (hx). The mean score in the clinical population is also provided; note that this does not necessarily represent a threshold for clinical significance. Finally, the Cronbach’s alpha, representing the internal consistency of the measure is provided, as reported in the cited work. | | | |

**Supplemental Table 4: Model 2 Results - POMP ~ Measure + IQ Composite + Participant Age + (1|WS-CG Pair)**

| **Term** | **B** | **SE** | **t-statistic** | **DOF** | **p-value** |
| --- | --- | --- | --- | --- | --- |
| *(Intercept)* | 0.1086 | 0.0864 | 1.2570 | 20 | 0.2228 |
| *GPTS-A* | 0.0062 | 0.0432 | 0.1436 | 288 | 0.8858 |
| *GPTS-B* | 0.0055 | 0.0432 | 0.1284 | 288 | 0.8978 |
| *PDI Endorse* | 0.0289 | 0.0432 | 0.6688 | 288 | 0.5041 |
| *PTQ Total* | 0.0480 | 0.0432 | 1.1121 | 288 | 0.2669 |
| *DERS Total* | -0.0151 | 0.0432 | -0.3507 | 288 | 0.7260 |
| *LSAS Endorse* | -0.0475 | 0.0432 | -1.0986 | 288 | 0.2728 |
| *LSAS Avoid* | -0.0299 | 0.0432 | -0.6930 | 288 | 0.4888 |
| *DWQ Worry* | -0.0010 | 0.0432 | -0.0236 | 288 | 0.9811 |
| *DWQ Paranoid* | -0.0220 | 0.0432 | -0.5104 | 288 | 0.6101 |
| *DWQ Total* | -0.0203 | 0.0432 | -0.4698 | 288 | 0.6387 |
| *BCIS-SR* | -0.0036 | 0.0432 | -0.0845 | 288 | 0.9327 |
| *BCIS-SC* | -0.0270 | 0.0432 | -0.6253 | 288 | 0.5322 |
| *LSNS Total* | 0.0191 | 0.0432 | 0.4428 | 288 | 0.6582 |
| *DACOBS JTC* | 0.0168 | 0.0432 | 0.3887 | 288 | 0.6977 |
| *DACOBS BIF* | 0.0328 | 0.0432 | 0.7606 | 288 | 0.4475 |
| *DACOBS SOC* | -0.0065 | 0.0432 | -0.1521 | 288 | 0.8791 |
| *IQ COMPOSITE* | -0.0015 | 0.0009 | -1.5946 | 16 | 0.1303 |
| *WS_AGE* | 0.0050 | 0.0022 | 2.3058 | 16 | 0.0348 |
| **Supplemental Table 4.** The results of the linear modeling of mixed effects (measure, composite IQ, and age) on percent of maximum possible difference (POMP) are shown. This includes the contrast estimate (B), standard error (SE), t-statistic, degrees of freedom (DOF) and p-value for each effect, as well as for the individual levels of measure. | | | | | |

**Supplementary Table 3 References**
